# Evaluation of Measles Surveillance System, Bono Region, Ghana

**DOI:** 10.1101/2023.09.29.23296365

**Authors:** Fidelis Zumah, Livingstone Asem, Amanda Debuo Der, Samuel Sackey

## Abstract

**Background:** Measles remains a global public health problem despite the availability of a safe and effective vaccine, it is one of the leading causes of childhood morbidity and mortality. Hence, this study assessed the performance of the measles surveillance system in the Bono Region of Ghana.

**Methods:** A descriptive evaluation study was employed using the updated Centers for Disease Control (CDC) guideline for evaluating public health surveillance systems. The data collection methods employed were observation checklists, questionnaires, and measles records review

**Results:** Out of the 12 districts, 83.3% had case definitions. Three-quarters had IDSR reporting forms while 66.7% could transport measles specimens for confirmation. The performance of the supportive functions of the system was sub-optimal. The study revealed that half of the districts were not able to meet most of the standards for the support function. Moreover, timeliness of weekly and monthly reports above 90% was found in the study to be 66.7% and 25% respectively, and just about half of the districts attained over 90% timeliness. Also, the measles surveillance system was simple, flexible, useful, and acceptable despite its low positive predictive value of 1.5%.

**Conclusion:** The results of the study suggest that the general performance of the measles surveillance system in the Bono Region was sub-optimal. The performance of the core functions of the surveillance system is considered to be satisfactory. About half of the districts performed satisfactorily for the supportive functions. The completeness and timeliness of the reports were also satisfactory. Also, the Measles surveillance system was simple, flexible, useful, and quite acceptable despite its low positive predictive value. There is a need for capacity building on measles surveillance for surveillance officers and community volunteers, effective supportive supervision, and effective communication at all levels to improve the activities of the surveillance system and ultimately eliminate measles.

## Background

Measles is an acute viral infection and a leading cause of childhood morbidity and mortality globally [1]. It is spread mainly through respiratory droplets or airborne spray to the mucous membrane in the upper respiratory tract [2]. However, after the introduction of routine measles vaccination as part of the Expanded Programme on Immunization in most developing countries including Ghana in the 1980s, global measles mortality has reduced [3]. Surveillance is the systematic, continuous collection, analysis, and interpretation of health-related data, which is needed for the planning, implementation, and evaluation of public health practice [4]. This has been operational in Ghana for measles and other priority diseases. It is meant to reduce morbidity and mortality and improve measles vaccination coverage to eliminate the disease in the country. The measles vaccine is one of the best investments in public health as it saved 23.2 million lives between 2000 and 2018 [5,6]. However, measles disease surveillance is a critical component of measles control and elimination efforts and is used in the assessment of progress and in making adjustments to programmes as required [7]. Developed countries such as the United States, Australia, and Japan have extensive measles surveillance systems that include both laboratory-based and clinical surveillance components whereby measles data are collected and analyzed from a variety of sources including laboratories, hospitals, general practitioners, and local health authorities. The insight from the data is used to monitor the spread of the disease and respond to outbreaks [8].

Measles surveillance is an ongoing monitoring of the occurrence of measles cases in a population. It is an important public health tool for identifying outbreaks of measles and for implementing control measures to prevent the spread of the disease [9]. In Ghana, the measles surveillance system uses a combination of active and passive surveillance methods which is similar to the measles surveillance system in Canada. The active surveillance method involves trained staff actively seeking out and collecting data on cases of measles using standardized reporting forms while the passive surveillance includes collecting data on measles cases that are reported to the surveillance system through other means such as media reports [10].

Since Ghana is in the measles elimination phase, the objective of measles surveillance is to detect outbreaks of fever with rash illness promptly and immediate case-based reporting of cases and deaths for routine surveillance and outbreaks; confirm the first five cases of suspected measles in a health facility per week with a laboratory test. The surveillance system also seeks to: strengthen the capacity of health systems to conduct effective surveillance activities; integrate multiple surveillance systems so that forms, personnel and resources can be used more efficiently and effectively; improve the use of information for decision-making; improve the flow of surveillance information between and within levels of the health system; improve laboratory capacity and involvement in confirmation of pathogens and monitoring of drug sensitivity; strengthen the involvement of laboratory personnel in epidemiological surveillance; increase the involvement of clinicians in the surveillance system; emphasize community participation in detection and response to public health problems [11].

Despite the remarkable efforts in measles vaccination coverage over the years towards its elimination in Ghana by 2020 under the Global Vaccine Action Plan, the country did not achieve the 2020 elimination milestones as 88 cases were reported in 2020 with a relatively higher, 1,274 number of cases registered in 2019 [12]. That is; 53% of the total cases reported in the last 2 years were reported in 2019 within seven months (January-August). The This could be because Ghana’s measles vaccine coverage is currently below the optimum threshold [12]. This made it difficult for Ghana to meet the 2020 measles elimination target set by the WHO. It is therefore imperative to evaluate the measles surveillance system to ensure that it can identify and confirm cases, and identify outbreaks [12]. This will stimulate further research in evaluating the performance of other communicable and non-communicable disease surveillance systems in the Bono Region and other regions in Ghana. It could contribute directly to the achievement of sustainable development goal 3 which calls for good health and well-being for all [13].

To the best of my knowledge, no studies have been conducted on the performance of the measles surveillance system in the Bono Region. For this reason, this study evaluated the performance of the measles surveillance system to provide useful information to facilitate the elimination of the disease in the Bono region and Ghana as a whole. The core and supportive activities including the attributes were also assessed.

## Materials and methods

### Study Area

The study was conducted in the Bono region of Ghana. It has a population of about 1,146,220 people and an annual population growth rate of 2.3% [14]. There are 12 districts in the region with Sunyani as the administrative capital with over 401 public and private healthcare facilities distributed across the region. Measles surveillance is conducted in all 12 district health directorates. There have been sporadic outbreaks of measles from 2015 to 2019 in some of the districts including Sampa, Banda, Sunyani West, Dormaa Municipal, Drobo, Tain, and Nkrankwanta with a total of 11 confirmed cases with no recorded mortality [15].

### Study design

A descriptive cross-sectional evaluation study was employed in this study using the Centers for Disease Control and Prevention (CDC) updated guidelines for evaluating public health surveillance systems in 2001 [5]. This design was used because it provided a snapshot of the measles surveillance system at a specific point in time. Also, it was appropriate for identifying the strengths and weaknesses of the measles surveillance system based on the performance of its core functions, supportive functions, and attributes and for making recommendations for improvement.

### Study population

The study population was all health workers in the District Health Management Team (DHMT) play a key role in measles surveillance activities. A total of 65 surveillance officers (34, disease control officers, and 28, field technicians) responded to the questionnaire.

### Inclusion and Exclusion Criteria

The study included all districts and their surveillance officers within the Bono Region who were willing to participate in the study. However, surveillance officers who were on their annual leave during the period of data collection were excluded. In addition, surveillance officers who were unwilling to participate in the study were excluded. Also, there was no incomplete filled questionnaire or checklist that was excluded.

### Sampling method/procedure

All 12 districts in the Bono Region were included in the study because measles surveillance was ongoing in each of them. The non-probability (purposive sampling) technique was used to select the surveillance officers (disease control officers and field technicians) in the Bono Region due to their involvement and participation in surveillance activities. This sampling technique was appropriate because it allowed surveillance officers who are knowledgeable and experienced with measles surveillance to give their unique insights.

### Data collection techniques/methods and tools

A structured questionnaire and an observation checklist adapted using the Centers for Disease Control and Prevention (CDC) updated guidelines for evaluating public health surveillance system was used. The data collection methods employed were an observation checklist and an interviewer-administered structured questionnaire. This is because the observation checklist and structured questionnaire as different data collection tools were used to complement each other to overcome the limitations of individual tools in assessing the measles surveillance system. The observation checklist was used to assess the performance of the core functions, and supportive functions as well as the data quality aspect of the attributes (completeness and timeliness) of the surveillance system. The checklist was issued to each of the districts. The performance of the measles surveillance attributes was assessed using a structured questionnaire. Measles data extracted from the DHIMS 2 from 2010 to 2019 was used to determine the trend of reported measles cases and immunization coverage in the Bono Region.

### Training of Field Staff and Pre-testing

Four field assistants with a minimum of first degree qualifications were trained for a day to help in the data collection. The data collection tools were pre-tested in Kenyasi District in the Ahafo Region to ensure the internal validity and reliability of tools and the necessary corrections were made. The Kenyasi District was chosen for the pre-testing because it had similar surveillance operation activities as compared to the Bono Region. Also, it was easier to obtain the necessary permission and approval for pretesting in the Kenyasi District.

### Data processing and management

The collected data were screened to ensure that all the needed information was correctly captured after every fieldwork. The data collection tools were given unique identifiers for easy identification and verification. The core, supportive surveillance functions and the surveillance attributes (completeness, timeliness, simplicity, flexibility, acceptability, usefulness and stability) sessions on both the checklist and the structured questionnaires were entered into EpiData version 4.0 while reported measles cases and immunization coverage data from 2010 to 2019 were also extracted to Microsoft Excel 2013.

### Data analysis/ statistical methods

The data were exported to STATA IC version 15.0 by StataCorp LLC for analysis. Descriptive statistics were used and frequency tables were used to display the results of the core and supportive functions of the surveillance system. For the attributes of the surveillance system, the completeness of case-based, weekly and monthly reports of the districts was categorized into 90%-94% and 95% and above. Also, the timeliness of the weekly and monthly reports of the districts was categorized into three (> 80%, 80% to 90%, and 90% and above). Also, the questionnaire on the surveillance system attributes was answered by each of the 65 surveillance officers whereby 1=20%, 2=40%, 3=60%, 4=80%, and 5=100%. The average score for each attribute was determined and further classified into three (3) categories, >60%, 60% to 79%, and 80% and above accordingly. The predictive positive value (PPV) was calculated in percentage using the extracted data from 2010-2019 in the DHIMS 2. Thus; PPV= True Positive/ (True Positive + False Positive) whereby True Positive=Laboratory confirmed measles cases, False Positive= (Suspected measles cases – Laboratory confirmed measles cases)

### Ethical Issues

Ethical approval was sought from the Ghana Health Service Ethics Review Committee (GHS-ERC) (GHS-ERC 024/06/20). Local permission and approval for the study were obtained from the Bono Regional Health Directorate to use their secondary source data (measles case-based reports and data from the DHIMS 2). Also, permission was sought from district health directorates, as well as district surveillance officers coordinating the surveillance activities at the various districts. Data was protected and only researchers had access to it. Participating in the study was voluntary and the purpose of the study was clearly explained to the health workers so they decide whether to consent or not. Only those who provided consent were included in the study. There were no direct benefits for participants. Surveillance officers were assured that under no condition whatsoever will their names or any other contacts be linked to the data analysis and dissemination of the findings of the study.

## Results

### Trend of reported measles cases and immunization coverage, 2010-2019, Bono Region

Figure 1 shows suspected and laboratory confined Measles cases recorded from 2010 to 2019 in the Bono Region. As MCV2 coverage for 18-59 months drops from 84.7% to 61.5% in the year 2010-2013, the number of suspected Measles cases detected by the surveillance system increased from 19 to 77 as the number of laboratory-confirmed cases shot up from 0 from 7 as well within the same period. The number of suspected and laboratory-confirmed measles continue to increase from 2014 to 2017 with a marginal drop in MCV2 coverage over the same period. The year 2018 recorded the highest, 222 suspected cases with a drop, of 184 cases in 2019. However, the highest MCV2 immunization coverage of above 90% was seen in 2019 with 4 laboratory-confirmed cases. Generally, from 2010 to 2017, there was a reduction in MCV2 immunization coverage from 84.7% to 80.4% with a corresponding increase in both suspected and laboratory-confirmed cases. Despite the high number of suspected and laboratory-confirmed cases, no measles-related death was recorded in the Bono Region.

**Figure 1:**
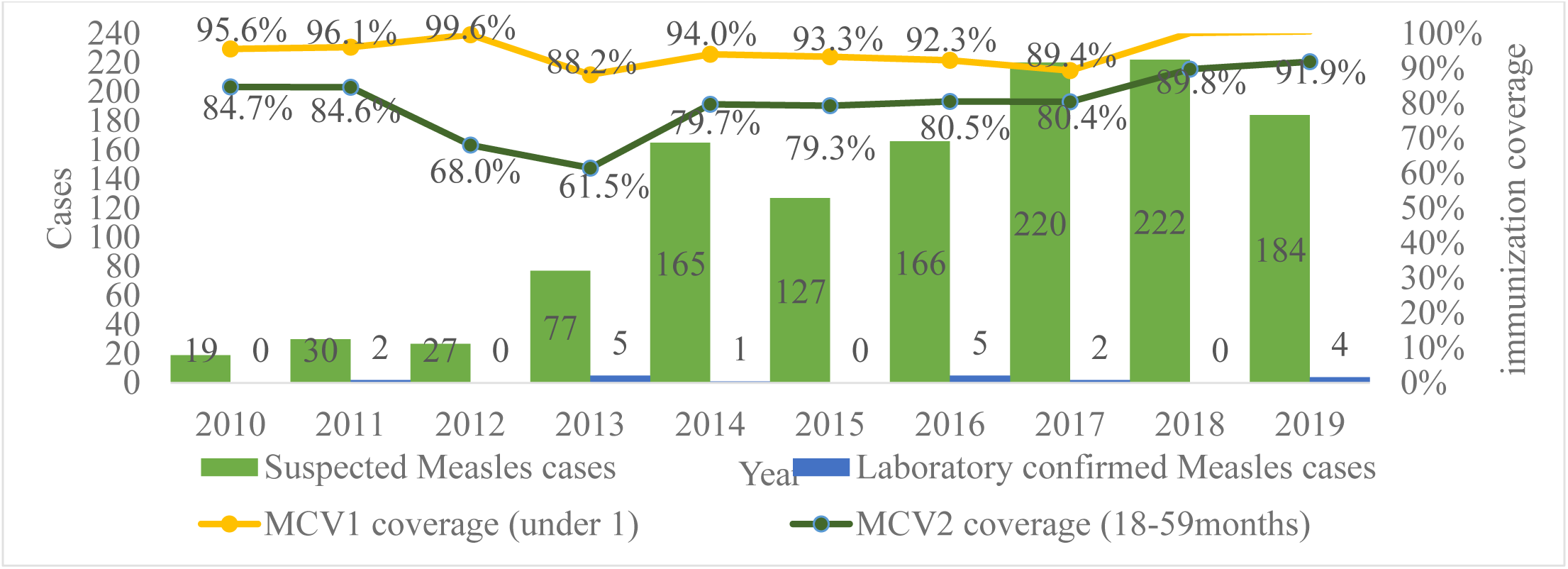
Trend of reported measles cases and immunization coverage, 2010-2019, Bono Region.

### Case reporting: Means of reporting suspected measles cases

Out of the 12 districts interviewed on how suspected measles cases were being reported to the Bono Regional Surveillance and Disease Control Unit, almost all (91.7%) said through a telephone call.

### Performance of core functions of the surveillance system, Bono Region

All 12 districts in the Bono Region were assessed on the performance of the core functions of the measles surveillance system. On case detection, standard case definitions were available in the majority (83.3%) of the districts. Most of the districts (75%) had IDSR reporting forms available in their files and they did experience a shortage in the past 6 months. Also, the majority (66.7%) of the districts had means of transporting their measles samples to the region. However, 75% had no guidelines for specimen collection, handling, and transportation. Additionally, on surveillance data analysis, 83.3%, 66.7%, and 75% of the districts did not describe their measles surveillance data by person, time as well as perform trend analysis respectively. Conversely, most (66.7%) described their data by place using spot maps.

Moreover, most (66.7%) districts had no appropriate denominator available for calculating risk population. Nevertheless, 66.7% had an action threshold for measles, and 75% had their action thresholds correctly stated. Regarding epidemic preparedness, the majority (91.7%) of the districts had no epidemic preparedness plan for measles surveillance. Almost all districts (91.7%) had access to funds from the IDSR budget for emergency response. Furthermore, there was the existence of a functional epidemic management committee in 66.7% and all the districts had rapid response teams (RRTs). Concerning feedback, three-quarters of the districts never received any feedback on measles surveillance activities despite their active participation. In the same vein, three-quarters (75%) of the districts had no available written report(s) that is regularly produced to disseminate surveillance data in their respective districts (Table 1).

**Table 1:** Observed Evidence of Performance of core functions of the surveillance system, Bono Region.

| **Surveillance Core Functions** | **Number (Percentage)**<br>**N (%)** |
| --- | --- |
| **Case Detection** |  |
| Have standard definition for measles | 10(83.3) |
| Case Reporting |  |
| Availability of IDSR reporting forms | 9(75.0) |
| Shortage of IDSR reporting forms during the last 6 months | 3(25.0) |
| **Case Confirmation** |  |
| Capacity to transport measles specimen to higher level laboratory | 8(66.7) |
| Have guidelines for specimen collection, handling and transportation | 3(25.0) |
| **Data Analysis** |  |
| The district described data by person | 2(16.7) |
| The district described data by time | 4(33.3) |
| The district described data by place | 8(66.7) |
| The district performed trend measles data analysis | 3(25.0) |
| The district had appropriate denominators (observe availability of population) | 4(33.3) |
| The district had an action threshold for measles | 8(66.7) |
| Districts with Measles threshold stated correctly, Available (n=8) | 6(75.0) |
| **Epidemic Preparedness** |  |
| A written epidemic preparedness plan | 1(8.3) |
| Budget line or access to funds for emergency response | 11(91.7) |
| Functional epidemic management committee | 8(66.7) |
| Districts had a rapid response team | 12(100.0) |
| **Feedback** |  |
| District receive feedback on measles from the next level | 3(25.0) |
| Presence of a written report that is regularly produced to disseminate surveillance data | 3(25.0) |

### Performance of supportive functions of Measles surveillance system, Bono Region

As shown in Table 2, about supervision, the majority (75%) of the districts had been supervised generally on surveillance including measles in the past 6 months. But, 58.3% had not done supervisory visits to their sub-districts and facilities. Also, 66.7% had not been trained on the surveillance system. Similarly, 66.7% had not conducted any refresher training at the sub-district and facility levels in the past year. Moreover, almost all (91.7%) had no bicycles for community surveillance volunteers but 66.7% had motorbikes for surveillance activities. Additionally, 75.0% had no protective materials respectively. To add, 91.7% had computers, however, none had a statistical package for surveillance data management. Also, 50.0% and 66.7% had internet services and stationery respectively available for general IDSR activities including measles.

**Table 2:** Observed Evidence of Performance of supportive functions of Measles surveillance system, Bono Region.

| Supportive Functions | Number (Percentage)<br>N (%) |
| --- | --- |
| District has been supervised on surveillance in the last 6 months | 9 (75.0) |
| Made supervisory visits in the 6 months | 5(41.7) |
| Trained in disease surveillance in the last 1 year | 4(33.3) |
| Conducted surveillance training in the last 1 year | 4(33.3) |
| Availability of bicycles for surveillance activities | 1(8.3) |
| Availability of motorcycle(s) for surveillance activities | 8(66.7) |
| Availability of vehicles(s) for surveillance activities | 5(41.7) |
| Availability of protective materials | 3(25.0) |
| Availability of computer for surveillance activities | 11(91.7) |
| Unavailability of statistical package | 12(100.0) |
| Availability of internet services | 6(50.0) |
| Availability of stationery | 8(66.7) |

### Performance of Measles surveillance Attributes, Bono Region

From the records reviewed of all 12 districts, only 41.7% met the WHO benchmark of 95% and above completeness of compulsory fields filled on their case-based forms. Also, 66.7% and 58.3% had completeness scores of 95% and above for weekly and monthly reports respectively in the last three months. Further, regarding timelines of reports, 90% and the above score was recorded for 66.7% of the districts for the weekly reports, and less than 80% timeliness was seen for 41,7% of the districts. However, only a quarter met the WHO target of 90% and above timeliness in the past three months (Table 3).

**Table 3:** Measles surveillance data completeness and timeliness, Bono Region.

| **Surveillance Attributes: Data completeness and timeliness** | **Number (N=12)** | **Percentage (%)** |
| --- | --- | --- |
| **Percentage completeness of case-based forms** |  |  |
| 90% - 94% | 7 | 58.3 |
| 95% and above | 5 | 41.7 |
| **Percentage completeness of weekly reports in the last three months** |  |  |
| 90% - 94% | 4 | 33.3 |
| 95% and above | 8 | 66.7 |
| **Percentage completeness of monthly reports in the last three months** |  |  |
| 90% - 94% | 5 | 41.7 |
| 95% and above | 7 | 58.3 |
| **Percentage timeliness of weekly reports in the last three months** |  |  |
| Less than 80% | 2 | 16.7 |
| 80% - 89% | 2 | 16.7 |
| 90% and above | 8 | 66.7 |
| **Percentage timeliness of monthly reports in the last three months** |  |  |
| Less than 80% | 5 | 41.7 |
| 80% - 89% | 4 | 33.3 |
| 90% and above | 3 | 25.0 |

On the performance of the other measles surveillance attributes, 65 surveillance officers (Disease Control officers and field technicians) were interviewed to rate the surveillance attributes in their respective districts. Of these, more than half (53.9%) rated the system to be simple and nearly half (47.7%) said the system was somewhat flexible. Furthermore, of the 65 participants, only 23.1% said the system was acceptable. Finally, regarding usefulness, more than half of the participants (55.4%) rated the measles surveillance system to be somewhat useful (Table 4).

**Table 4:** Performance of Measles surveillance Attributes, Bono Region.

| **Measles surveillance attributes** | **Number (N=65)** | **Percentage (%)** |
| --- | --- | --- |
| **Simplicity of the surveillance system** |  |  |
| Not Simple | 4 | 6.2 |
| Somewhat Simple | 26 | 40.0 |
| Simple | 35 | 53.9 |
| **Flexibility of the surveillance system** |  |  |
| Not flexible | 7 | 10.8 |
| Somewhat flexible | 31 | 47.7 |
| Flexible | 27 | 41.5 |
| **Acceptability of the surveillance system** |  |  |
| Not acceptable | 28 | 43.1 |
| Somewhat acceptable | 22 | 33.9 |
| Acceptable | 15 | 23.1 |
| **Usefulness of the surveillance system** |  |  |
| Not useful | 11 | 16.9 |
| Somewhat useful | 36 | 55.4 |
| Useful | 18 | 27.7 |

## Discussion

### Performance of core measles surveillance systems functions

Standard case definitions were available in most of the districts in this current study. Similar findings were revealed by studies conducted in India and Kenya [16,17]. The availability of standard case definitions indicates that most measles diagnoses are likely to be correct if used effectively. Also, two-thirds of the districts had both weekly and monthly IDSR reporting forms field. Contrasting results were, however, revealed by a study conducted in India [16]. The findings from this current study showed that surveillance activities are always ongoing and efforts are made by healthcare providers to report the activities of the surveillance. This is necessary for tracking the progress toward the elimination of measles. In addition, most of the districts could transport the specimen to the region for onward transportation to the central laboratory (Public Health Reference Laboratory, Korle-Bu). Comparatively, similar results were shown in a study conducted in India [16]. The capacity of the districts to transport samples to the reference laboratory is an indication of a commendable relationship between the districts and the reference laboratory and also the commitment to confirm suspected cases and the performance of the whole surveillance system.

Furthermore, this study bared weak data analysis at all districts, and only a few analyzed data by person, time, and trend. Elsewhere in India and Nigeria, only a few districts analyzed their surveillance data [16, 18]. The weak data analysis could be attributed to inadequate technical officers, limited training on surveillance measles, minimal supervision, inadequate data analysis capacity, and lack of data management resources. This hinders the effective detection of outbreaks and the use of data for planning and decision-making [16, 19].

Additionally, two-thirds of the districts had functional epidemic management committees. Similar results were found in a study in India [16]. The presence of functional epidemic management could mean that they are always ready to respond in case of an outbreak. However, there is always a gap between the presence of such committees and their response ability [20]. To add, the feedback system at the district level was weak. This was evident as only one-fourth of the districts said they receive feedback on the measles surveillance activities from the region and had verifiable evidence. This may be due to delays in receiving feedback on samples collected from the central laboratory. This conforms to findings in a study conducted in Tanzania [21] and a multi-country study conducted in Ghana and Tanzania [22]. Contrary results were, however, reported in India [16]. Regardless of the setting, feedback is very important in communication and could be considered essential for the performance of the measles surveillance system as such information is used for making decisions relating to treatment and contact tracing.

### Performance of supportive functions of the measles surveillance system in the Bono Region

Integrated disease surveillance, laboratory, and outbreak investigation guidelines support surveillance activities [23]. This study found that the majority of the districts did not have guidelines for specimen collection, handling, and transportation. However, contrasting results were reported in a study conducted in Ethiopia [24]. The unavailability of guidelines in this current study setting could be a setback in measles surveillance as it is one of the diseases earmarked for elimination in the IDSR even though Ghana has been reported to have made substantial progress [25]. Moreover, the majority of the districts had neither trained nor conducted any refresher training at the sub-district and facility level in the past year. The training of staff enables them to be updated on changes and new practices in the measles surveillance system [23]. This is in contrast to a study in India [16]. There could be no funds for training in the current study population. However, this tends to make staff obsolete to current practices in the surveillance system. Further, three-quarters of the districts had been supervised on measles surveillance. Conversely, a study conducted in Ethiopia reported otherwise [24]. Supportive supervision for measles surveillance ensures that the required logistics are in place, builds the capacity of staff, and ensures timely implementation of planned surveillance activities [23].

Moreover, most of the districts in this current study had no means of transportation to conduct measles surveillance activities. Similar findings were obtained in India [16]. Means of transport is very important for the successful conduct of surveillance activities [23]. The unavailability of transportation could be a huge hindrance to the surveillance system. To add, almost all the districts had computers, however, none had a statistical package for surveillance data management. Contrasting findings were reported by a study conducted in India [16]. The importance of data management in the measles surveillance system cannot be overemphasized as it has significant effects on decision-making. Also, a measles surveillance system can be undermined if there are no resources to capture and analyze data as it will give opportunity for abrupt outbreaks and a hindrance to the goal of measles elimination.

### Performance of the Attributes of the Measles Surveillance System in the Bono Region

#### Completeness

In this study, the majority of the districts had 90% - 94% completeness of case-based forms. This finding differs from the results reported in Taiwan [26]. However, a multi-country study conducted in Ghana and Tanzania confirmed these findings [22]. Additionally, this finding is consistent with the 2019 reporting rate completeness of IDSR reports in DHIMS2 [27].

#### Timeliness

Also, only a quarter of the districts met the national target of 90% timeliness. This is consistent with findings from a study conducted in Northern Ghana [28]. However, it slightly differs from the timeliness of IDRS reports in DHIMS 2 [27]. As measles is earmarked for elimination, the acceptable timeliness for reporting is 100%. This is to ensure that suspected cases are promptly investigated and guide the program managers to make time-bound decisions.

#### Simplicity

The measles surveillance system in the Bono region was found to be simple as the case definitions for the identification of suspected cases are easy to understand and can easily be translated into the local language. Also, most surveillance officers actively participate in surveillance activities. Further, home visits or phone calls are usually done at the facility and community level to obtain information about a case. Similar findings were reported by a study conducted in Taiwan [26]. This demonstrates the easy way of staying in touch with the community to facilitate effective surveillance activities.

#### Flexibility

It was also found that about half of the surveillance officers rated the system to be somewhat flexible. This means that the possibilities of changes in activities could be easily accommodated in the measles surveillance system. This is similar to findings from a study conducted in Southeast Ethiopia [29]. This highlights the importance of a flexible surveillance system, especially in the elimination of measles.

#### Acceptability

Of the surveillance officers, 57% rated the measles surveillance system as acceptable. Acceptability is often determined by the willingness of the surveillance officers to effectively implement the system through active participation in its activities. This is expected to be 100% and rated using attributes such as completeness of case-based report forms and timeliness of data reporting [29]. This could be due to the challenges in engaging clinicians in measles surveillance and the loss of interest in community-based health volunteers.

#### Usefulness

The data generated by the measles surveillance system were said to be somewhat useful which is contrary to findings from a study conducted in Uganda [30]. This implies that the data on measles surveillance in the Bono Region of Ghana can be used for planning and decision-making. Also, it is an indication that the data generated in the districts are reliable.

#### Positive Predictive Value

The positive predictive value (PPV) of the measles surveillance system was found to be low (1.5%). This implies that for all the suspected measles cases that the surveillance system detects, only 1.5% of them were true events/confirmed cases. This means that the system has low accuracy in detecting significant events (true measles cases), and a high rate of false positives which could lead to unnecessary investigations and waste of resources. This could also lead to a lack of confidence in the system and its ability to accurately detect measles cases. On the contrary, a low PPV of 1.5% may be attributed to the low prevalence of the disease in the region. There could also be a possibility of increased vigilance for measles leading to the reporting of vaccination-associated rash resulting in low true IgM positive results. Similar results were discovered in a study conducted in Ontario [31]. Contrasting results were, however, reported in a study conducted in Tanzania [21]. Even though the surveillance system could be regarded as being sensitive due to the detection of a high number of suspected measles cases, its sensitivity could not be quantified because there was no data on the measles false negative parameter.

### Policy and Public Health Implications

The evaluation of the measles surveillance system is particularly necessary and of public health importance as the disease is targeted for elimination. The effective use of the available measles standard case definitions for the identification of suspected cases is paramount in ensuring accurate diagnosis and preventing the waste of resources for further investigation and contact tracing. The use of IDSR guidelines and report forms gives the region the precedence of tracking the progress of measles surveillance activities. To add, surveillance officers in the Bono region need to be abreast with data management and data analysis to always use the data obtained from their surveillance activities for decision-making and detecting outbreaks. The feedback system should be strengthened to facilitate effective surveillance information.

Completeness and timeliness of measles data are often expected to be 100% for prompt action. This was, however, not recorded in the Bono Region and could lead to unidentified events and delayed interventions. The simplicity and flexibility of the measles surveillance system are commendable as it makes surveillance activities easier for surveillance officers and community volunteers. Consequently, the low acceptability of the surveillance system poses a threat to all its activities. Surveillance officers and community volunteers could be motivated to actively participate in surveillance activities. This should be coupled with effective supportive supervision. Overall, the performance of the measles surveillance system in the Bono Region of Ghana will have negative effects on the achievement of SDG 3 if deliberate measures are not taken for improvement.

### Study limitations

Limited data for some years in some districts on suspected and confirmed measles cases and immunization coverage might have affected the findings of this study. Due to this, the data were inadequate to calculate the sensitivity of the surveillance system. Also, it is impossible to study the trends or changes in the measles surveillance system over time using a cross-sectional evaluation design, unlike a comparative study which could help provide a more comprehensive evaluation of the surveillance system and its effectiveness.

## Conclusion

The general core function performance of the measles surveillance system was considered to be satisfactory. Additionally, measles data analysis and feedback in most districts were weak and a major concern in the region. About half of the districts performed satisfactorily in the supportive surveillance functions. Further, the majority of the districts attained more than 95% report completeness, and just about half of the districts attained over 90% timeliness. Also, the Measles surveillance system was simple, flexible, useful, and quite acceptable despite its low PPV of 1.5%checklists. The system was however unstable as it depended highly on donor funding.

### Recommendations

With support from the national surveillance department, disease surveillance capacity-building training should be conducted at least twice a year for all health personnel in the region. Furthermore, supportive supervision of health facilities should be improved. Finally, the central laboratory should be urged to test suspected measles cases on time to reduce the lag time in case confirmation and feedback at all levels.

## Data Availability

All data produced in the present study are available upon reasonable request to the authors

